# A hybrid multi-scale model of COVID-19 transmission dynamics to assess the potential of non-pharmaceutical interventions

**DOI:** 10.1101/2020.04.05.20054460

**Authors:** Anass Bouchnita, Aissam Jebrane

## Abstract

Severe acute respiratory syndrome coronavirus 2 (SARS-CoV-2) is a novel coronavirus that emerged in Wuhan, China in December 2019. It has caused a global outbreak which represents a major threat to global health. Public health resorted to non-pharmaceutical interventions such as social distancing and lockdown to slow down the spread of the pandemic. However, the effect of each of these measures remains hard to quantify. We design a multi-scale model that simulates the transmission dynamics of COVID-19. We describe the motion of individual agents using a social force model. Each agent can be either susceptible, infected, quarantined, immunized or deceased. The model considers both mechanisms of direct and indirect transmission. We parameterize the model to reproduce the early dynamics of disease spread in Italy. We show that panic situations increase the risk of infection transmission in crowds despite social distancing measures. Next, we reveal that pre-symptomatic transmission accelerates the onset of the exponential growth of cases. After that, we demonstrate that the persistence of SARS-CoV-2 on hard surfaces determines the number of cases reached during the peak of the epidemic. Then, we show that the restricted movement of the individuals flattens the epidemic curve. Finally, model predictions suggest that measures stricter than social distancing and lockdown were used to control the epidemic in Wuhan, China.

## 1. Introduction

Severe acute respiratory syndrome coronavirus 2 (SARS-CoV-2) is a highly transmissible coronavirus that has spread around the world causing an epidemic outbreak [1]. SARS-CoV-2 is a virus which can be transmitted to humans, in a similar way to other coronaviruses like SARS-CoV and Middle East Respiratory Syndrome (MERS)-CoV [2]. Human-to-human transmission occurs via respiratory droplets secreted by infected patients when they cough or sneeze. Some of these droplets can be directly be inhaled by individuals nearby. Others can contaminate neighbouring hard surfaces. As a result, a susceptible individual can contract the virus by touching a contaminated surface and then their face. This indirect mechanism of transmission is important in the case of SARS-CoV-2, because stable SARS-COV-2 can remain on some type of surfaces like plastic and stainless steel for a few days [3].

Infected individuals with coronavirus disease (COVID-19) do not develop symptoms immediately upon contracting the virus. Indeed, one of the main features of COVID-19 is the relatively important period of incubation. The median value for COVID-19 incubation period was estimated at 5.1 days [4]. However, a recent study suggested pre-symptomatic transmission of COVID-19 [5]. In other words, infected individuals start transmitting the disease a few hours or days before the end of the incubation period. This mode of ‘silent transmission’ could be a driver of the disease spread. SARS-CoV-2 causes moderate to severe clinical outcomes in 20 % of infected patients [6].

In the absence of effective vaccines or therapeutics, non-pharmaceutic interventions (NPIs) were adopted as a strategy to slow down the disease propagation. Some of these control measures aim to reduce the probability of infection transmission during direct contact. For example, by promoting hygiene practices and cough etiquette. Other measures, like social distancing and the closing of schools and workplaces, reduce the opportunities of human-to-human transmission. The rest of the interventions like the disinfection of public areas aim to reduce the chances of indirect transmission. While all of these measures slow down the propagation of the epidemic [7], the impact of each of these NPIs on the epidemic dynamics of COVID-19 remains hard to quantify.

Mathematical modelling can be used to gain a deeper understanding of the transmission dynamics of infectious diseases. Compartmental models provide a theoretical framework to study disease transmission in a population of individuals [8]. These compartmental models can be implemented using a wide range of modelling methods. The simplest form of compartmental models uses ordinary differential equations to describe population dynamics during an epidemic. Susceptible-infected-recovered (SIR) models are one of the most commonly used compartmental models [9]. They describe the evolution of the number of infected individuals in a closed population. A recently published SIR model was used to quantify the effect of quarantine on the spread of COVID-19 [10]. Another class of widely used models consider four population classes: susceptible-exposed-infectious-recovered (SEIR) models [11]. These models are most suitable to describe the spread of diseases with a long incubation period, which is the case of the COVID-19 outbreak. In this context, an SEIR was recently developed and used to conduct a data-driven analysis of COVID-19 spread [12]. Other recent studies used SEIR models to estimate the basic reproduction number of this pandemic [13, 14]. A more complex compartmental study was used to evaluate the capacity of the healthcare system in responding to COVID-19 propagation [15]. The impact of the control measures that were adopted to mitigate the COVID-19 outbreak in China was evaluated using another compartmental model [16]. While another study assessed the effect of unreported cases [17] on the outbreak dynamics. Compartmental models implemented using ODEs can be studied both analytically and numerically. Partial differential equations (PDEs) can be used to implement compartmental models to study the spatio-temporal dynamics of an epidemic. In this context, systems of reaction-diffusion equations can be used with to describe the spatial densities of populations such that the diffusion term represents the motility of individuals in a computational domain [18]. Numerical simulations of these models show the propagation of the disease in the form of travelling waves.

Multi-agent systems represent another framework which was extensively used to study the transmission dynamics of infectious diseases. The advantage of this approach is the possibility to accurately capture the interactions between individuals. In these models, each individual is represented as an agent that can communicate with the neighbouring individuals and with the environment. Sometimes, inter-agents communication is described using a dynamic network [19, 20]. Other times, the motion of agents is modelled and communications occur between neighbouring individuals. In this case, population movement and inter-agent communication can be described using cellular automata [21, 22]. social force models provide a more accurate description of pedestrian motion [23]. In these models, each agent aims to reach its desired velocity while keeping a certain distance from other agents. The effect of neighbouring individuals on the agent is described using a socio-psychological force. This approach can be used to describe the complex dynamics of crowds which involve several interacting individuals social force models can be applied to study the dynamics of infection spread in crowds and under realistic conditions. For example, a social force model was used to describe the infection propagation in airplanes [24]. Although more complex, agent-based models can yield results which are in a good agreement with simpler ODEs models [25].

Multi-scale modelling is a mechanism-based approach which aims to integrate the effects of regulatory mechanisms across multiple scales of space and time [26]. Hybrid discrete-continuous model benefits from the advantages of both the agent-based framework and the equation-based representation [27]. These models were developed to describe biological systems [28, 29]. In multi-scale models, Infection dynamics are represented at the host and the population levels [30]. The advantage of this approach is the possibility to integrate available data and knowledge across multiple scales of description. In this context, collected data was used to calibrate several multi-scale models of the foot-and-mouth epidemic [31]. Multi-scale models provide a rich description of the infection dynamics at the individual and environment levels than multi-agent models [32].

As of April 27, 2020, SARS-CoV-2 is a newly discovered coronavirus. The available data and knowledge about COVID-19 epidemics remain insufficient. As a result, it is hard to quantify the effectiveness of each non-pharmaceutical intervention in slowing down the spread of the pandemic. To gain a better understanding of the epidemic features of COVID-19, we develop a hybrid multi-scale model that describes the transmission dynamics of this disease at both the individual and population levels. We represent individuals as agents that move in a square computational domain. Their displacement is described using a social force model. Infective agents can transmit the virus through direct contact with susceptible individuals. They can also contaminate the neighbouring surfaces by secreting droplets which contain the virus. The concentration of stable SARS-CoV-2 on hard surfaces is described with a partial differential equation. Infected individuals do not develop symptoms until the end of the incubation period. However, they can start transmitting the virus before the onset of symptoms. The chances of survival of patients depend on their age and health conditions. We use the model to study the early transmission dynamics of COVID-19 in Italy, where control measures were relaxed. Next, we study the risk of virus transmission in crowds moving under panic. Then, we investigate the impact of pre-symptomatic transmission on the spread of the disease. After that, we quantify the effect of the effect of SARS-CoV-2 persistence on hard surfaces on the epidemic curve. Finally, we use the model to assess the potential of restricted population movement in slowing down the epidemic progression.

## 2. A hybrid model of COVID-19 transmission dynamics

In this section, we formulate a hybrid multi-scale model to describe the transmission dynamics of COVID-19. The model describes the spread of the infection in a closed population of 250 individuals, all potentially symptomatic, moving in random directions in a square section of 250 × 250 *m*^2^. Agents are introduced at random locations at the beginning of each simulation and the motion of individual agents is described using a social force model. We assume periodic boundary conditions to approximate systems with larger populations. A snapshot of numerical simulation using the model is provided in Figure 1. The model was previously applied to understand the impact of the NPIs adopted by Morocco in containing the COVID-19 epidemic [33].

**Figure 1:**
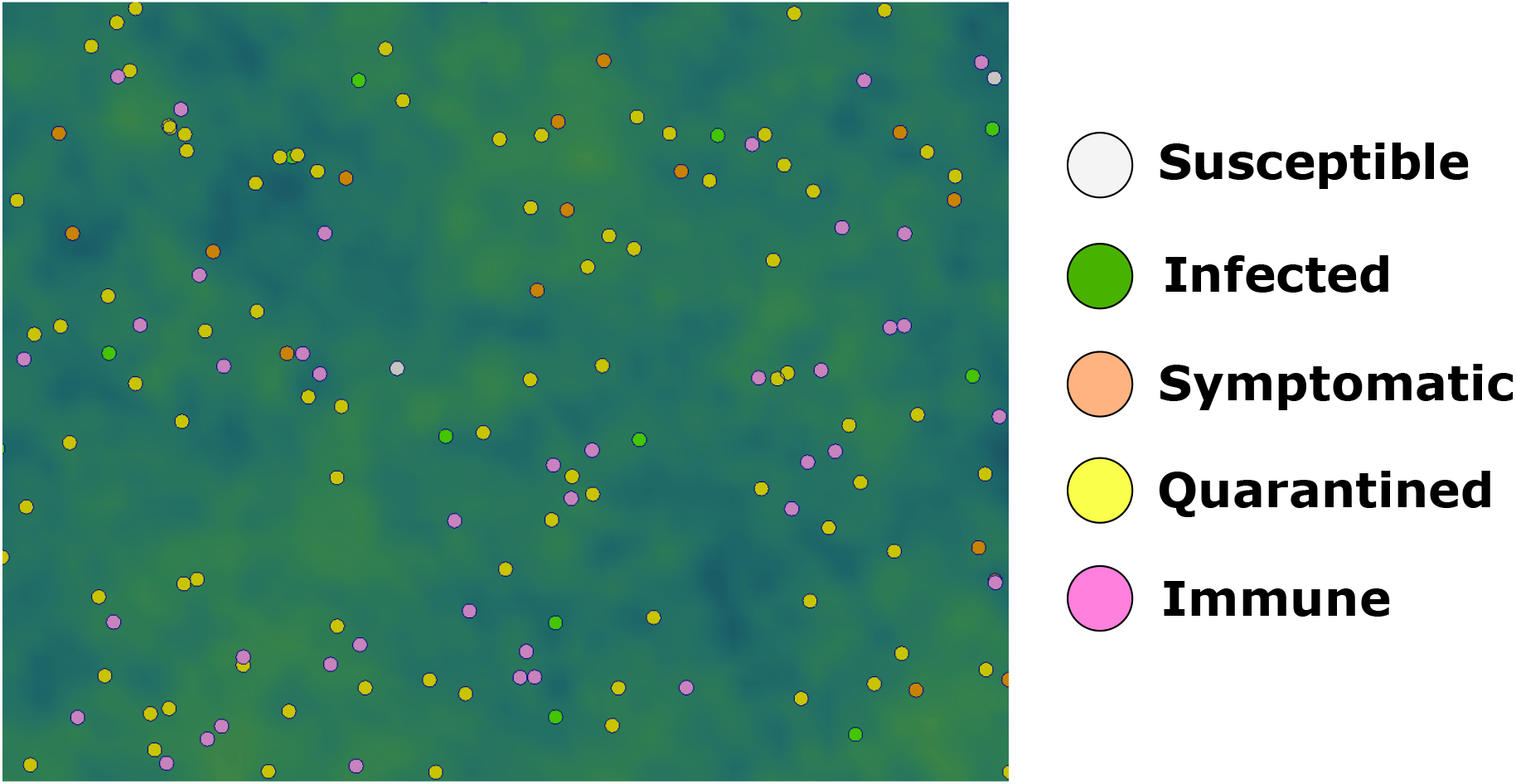
A snapshot of numerical simulation using the hybrid model. The size of agents is increased by six times to make the distinction of agents easier. Each type of agents is represented with a different color as shown in the legend. The green color gradient represents the concentration of the virus on contaminated surfaces.

We consider two modes of virus transmission. First, infective agents can transmit the disease to susceptible agents through direct contact. Second, they can contaminate neighbouring surfaces and subsequently cause the infection of other agents. We describe the concentration of the virus on hard surfaces using a partial differential equation. We consider that infected agents do not become symptomatic until the incubation period is over. Still, they can transmit the infection to susceptible agents before the end of this period. Infected agents become isolated a few days after the development of symptoms as they go into quarantine. They can either survive or die depending on their age and health conditions. The high-fidelity of the model is suitable for the study of NPIs and control measures which target both population movement and person-to-person disease transmission.

The objective of this study is not to give accurate predictions on the evolution of the epidemic. But rather to understand the effect of NPIs on the transmission dynamics of the disease. The present model is built on several assumptions. First, the role of children in disease transmission is not considered because most of the children that were infected with COVID-19 are asymptomatic [34]. Second, all the agents considered in the model are potentially symptomatic [35]. We do not consider asymptomatic individuals as we do not have sufficient data on the infection dynamics of these individuals. We restrict this study to symptomatic cases to compare with available epidemiological data. However, we consider pre-symptomatic transmission as it was demonstrated for COVID-19 by analyzing epidemiological data [5]. Third, we do not consider the effect of unreported cases on the dynamics of disease spread. Unreported cases could affect the transmission dynamics of the virus as they are not isolated from the rest of the population. The effect of unreported cases will not be considered in the present study to make the interpretation of results easier. Finally, we consider a relatively small computational domain with a reduced population of susceptible agents. Numerical simulations using this multi-scale model are computationally expensive. because we use a very small time step *(dt* = 10^−4^ h) to track all the contacts between agents. Therefore, stochastic noises are expected to be elevated in the obtained results, but the disease dynamics are expected to be the same as in larger systems.

### 2.1. Agent displacement

We study the dynamics of virus transmission in a population of agents that move in random directions. Each agent is represented by a circular particle of mass *m_i_* and radius *r_i_* as represented in Figure 2. We use modified Langevin equations to describe the velocity and displacement of each agent *i* [36]:

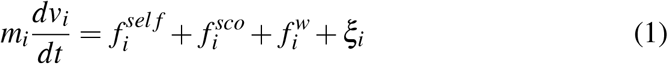

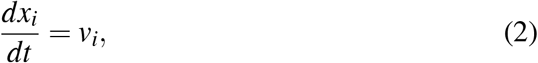

where *v*_1_ is the velocity of the pedestrian and *x_i_* is its displacement, 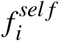 is the self-driven force (or the so-called acceleration force [23]) allowing the individual to reach his own destination is defined by:

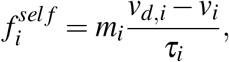

*v_d,i_* is the desired velocity at which the ith pedestrian wants to move. For normal conditions, the speed is chosen to be following a normal distribution with an average of 1.34 *m.s*^−1^ (≃ 5 *km/h)* and a standard deviation of 0.26 *m.s^−^*^1^, *τ_i_*. is a relaxation time, which specifies how long the pedestrian will take to recover his desired velocity after a contact or after a sudden change in their path. This parameter reflects the aggressiveness of the individual [37]. The social force 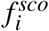 describes the tendency of agents to avoid contacts and keep a desired distance *d*_0_ between them in order to comply with social distancing measures [23]. It is the sum of repulsive forces exerted by neighbouring agents:

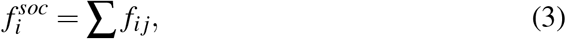

where 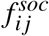 is given by:

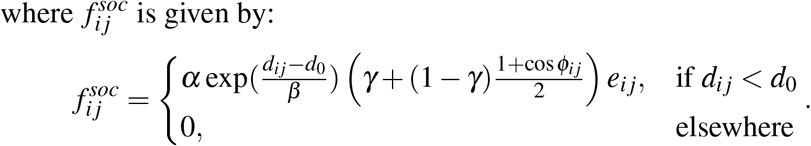

**Figure 2:**
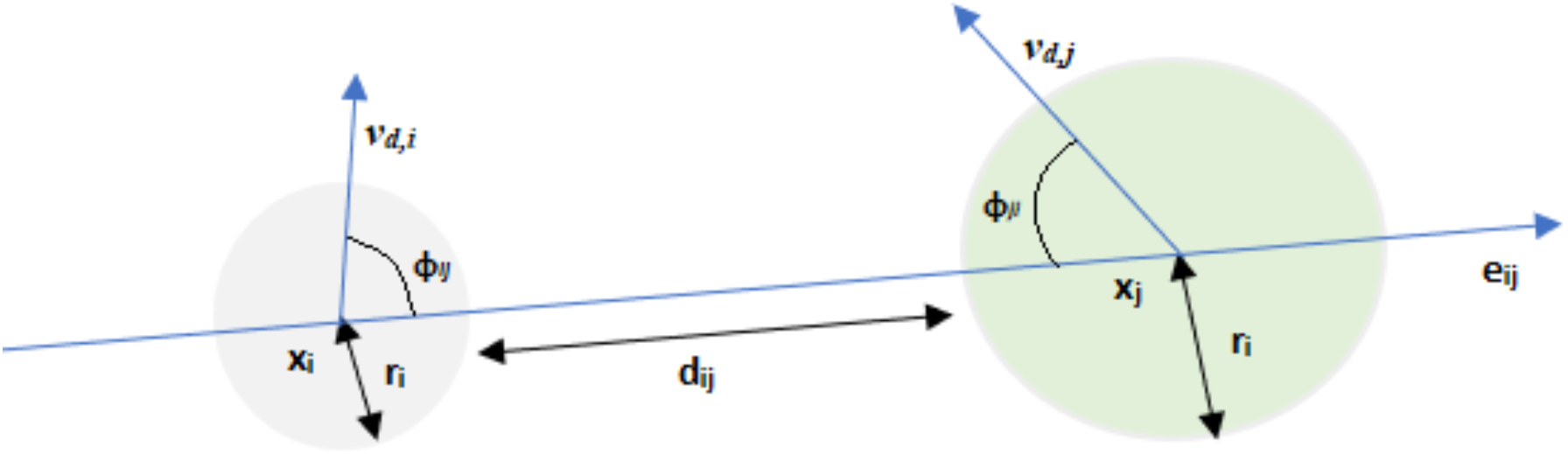
Schematic representation showing two pedesterian and the used notations in the description of the model.

Here, we have:

- *d_ij_*:= ||*x_i_ − x_j_* || − *(r_i_* + *r_j_*) is the distance between the two agents *i* and *j*.
- *d*_0_ is a desired distance that the individuals wish to keep between them, it can depend on the movement purposes of agents (leisure, evacuation, rush hour…) [36].
- *α* and *β* are two positive constants of the model representing the magnitude of the maximum socio-psychological force and its falloff length respectively.
- *γ* is a parameter taken such that 0 < *γ <* 1. It grows with the effect of interactions behind an individual, and *ϕ_ij_* is the angle between the desired velocity *v_d,i_* and the vector −*e_ij_*.

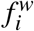 is also an exponential term that models interaction between pedestrian *i* and obstacles with defined such as introduced in [36], and *ξ_i_* is a small fluctuation force. The default values of parameters for the social force model are provided in Table 1.

**Table 1:** Default values for the parameters of the social force model taken from [37].

| Parameter | Symbol | Value | Unit |
| --- | --- | --- | --- |
| Walking speed | $\vert \vert v_d\vert \vert $ | N(1.34, 0.26) | $m.s^{-1}$ |
| Radius | $r_i$ | 0.23 | $m$ |
| Mass | $m_i$ | 83 | $kg$ |
| Relaxation time | $\tau_i$ | 0.5 | $s$ |
| Magnitude of socio-psychological force | $\alpha$ | 2000 | $kg.m.s^{-2}$ |
| Falloff length of socio-psychological force | $\beta$ | 0.08 | $m$ |

### 2.2. Modes of disease transmission

Infective individuals secrete droplets which contain SARS-CoV-2 when they cough or sneeze. These droplets transmit the virus if they get inhaled by other individuals directly. They can also contaminate neighbouring surfaces which can lead to the subsequent infection of susceptible persons. We consider both of these modes of transmission in our model. To describe direct contact transmission, we consider that each infective agent will probably transmit the virus to susceptible agents if the distance between them is less than one meter. Direct transmission depends on certain events such as sneezing, coughing, or handshaking. Thus, we can assume that person-to-person transmission follows a Bernoulli distribution. We consider that the susceptible agent has a probability (*p_d_*) of being infected upon contact with the contagious one. This probability can depend on the compliance with non-pharmaceutical measures such as the wearing of mask, handwashing, and cough etiquette.

Indirect transmission occurs when infective individuals contaminate the neighbouring hard surfaces. These individual transmit the virus to the surrounding surfaces by touch or through secreted droplets if they cough or sneeze. To simulate indirect contact, we first describe the normalized concentration of stable SARS-CoV-2 on hard surfaces:

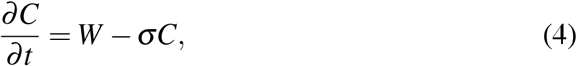

where *C* is the normalized concentration of deposited SARS-CoV-2 on hard surfaces, *W* is the averaged rate of SARS-CoV-2 secretion by contagious agents, *σ* is the decay rate of the virus. The daily probability of contracting the virus by indirect trnasmission is estimated as:

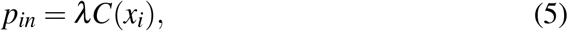

where *λ* is a positive constant taken smaller than one. Note that for each agent, indirect transmission can occur only once every day. It is evaluated at a random moment of the day for each agent.

We solve Equation 4 on a computational grid of 250 × 250 nodes using the Euler explicit scheme. Before solving this equation, we spread the production rate of secreted virus by infective individuals (*W*) to the computational grid using a radial basis function:

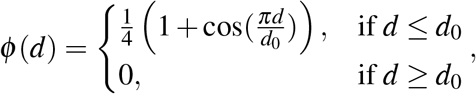

where *d* is the distance between the node and the center of the agent. *d*_0_ is the characteristic distance which describes the extent to which secreted droplets can infect the neighbouring surfaces. We take *d*_0_ = 2 *m*. To simulate indirect transmission, we evaluate the local concentration of stable SARS-CoV-2 at the center of susceptible agents. We compute this value using bilinear interpolation. Default values of transmission parameters are provided in Table 2.

**Table 2:** Default values for the parameters used in the submodel of disease transmission. The value of the decay rate was estimated such that the half-life time of stable SARS-CoV-2 on hard surfaces is taken equal to 4 *h* [3]. The probabilities of direct and indirect transmisstion were fitted to reproduce epidemiological data.

| Parameter | Symbol | Value | Unit |
| --- | --- | --- | --- |
| Secretion rate of SARS by infective agents | $W$ | 0.001 | $h^{-1}$ |
| Decay rate of SARS-CoV-2 | $\sigma$ | 0,1732 | $h^{-1}$ |
| Probability of direct transmission | $p_d$ | 0.04 | - |
| Parameter of indirect transmission per day | $\lambda$ | 0.015 | - |

### 2.3. The clinical course of infected patients

Infected individuals develop symptoms as soon as the period of incubation is elapsed. However, they become contagious and start spreading the disease a day before the end of this period [38]. The incubation period falls within the range of 2-14 days with a mean of 5.1 days. We use a log-normal distribution fitted using clinical data [4] to sample the values of the incubation period for each agent. After the development of mild symptoms, agents keep moving in the computational domain and transmitting the disease for a few days until the symptoms become more severe. Then, they become isolated and go into quarantine at home or in a hospital. We model this period of isolation by considering that the patient stops moving, transmitting the virus, and interacting with other agents. The outcome of this period will be either the death or the survival of the patient. If the agent survives, then it becomes immunized to infection and starts moving again. The considered COVID-19 epidemiological characteristics in the model are provided in Table 3.

**Table 3:**
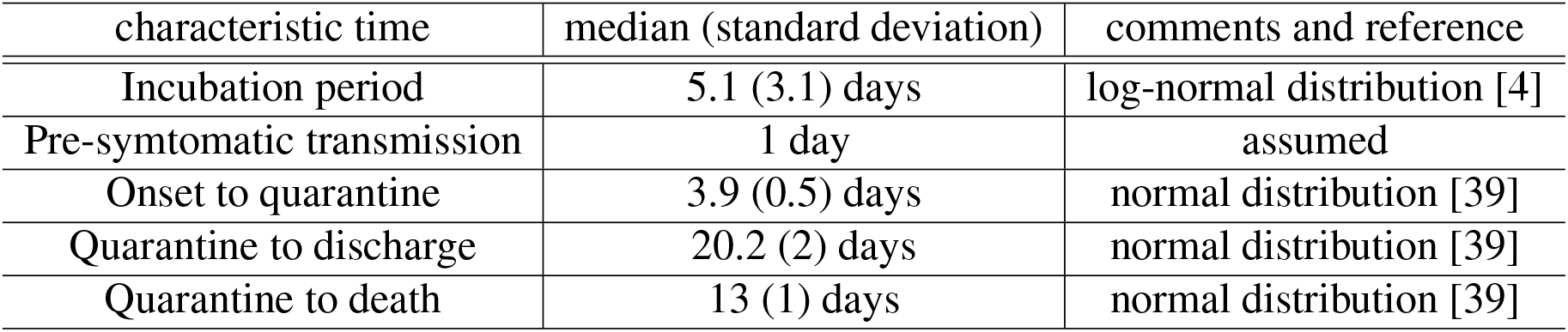
Values of the considered COVID-19 characteristics in the model.

| characteristic time | median (standard deviation) | comments and reference |
| --- | --- | --- |
| Incubation period | 5.1 (3.1) days | log-normal distribution [4] |
| Pre-symptomatic transmission | 1 day | assumed |
| Onset to quarantine | 3.9 (0.5) days | normal distribution [39] |
| Quarantine to discharge | 20.2 (2) days | normal distribution [39] |
| Quarantine to death | 13 (1) days | normal distribution [39] |

The mortality rate of each agent depends on the characteristics of the agent (age, risk factors) as described below. The median time from hospital admission to death is 13 days [39]. The median time from hospital admission to discharge for surviving patients is approximately 20.2 days. We have represented the clinical used course for infected patients in Figure 3.

**Figure 3:**
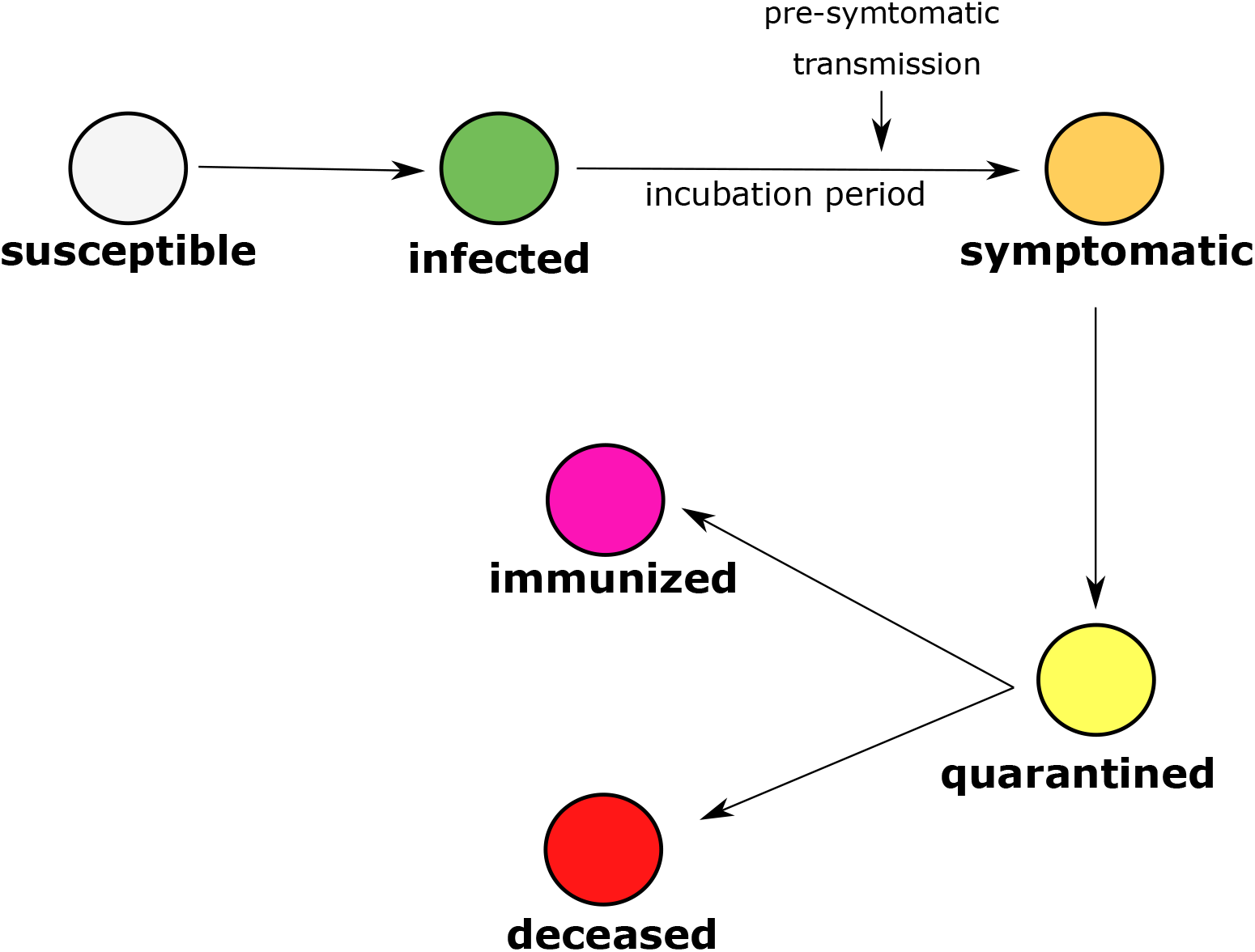
The clinical course of an infected patient in the model. The infected patient develops symptoms by the end of an incubation period. However, they can start transmitting the virus before the onset of symptoms. A few days after, patients become quarantined as their symptoms become more severe. Then, they can either die or survive and become immune depending on their age and health condition.

### 2.4. Demographic characteristics and mortality risk of infected agents

In our model, the mortality risk by COVID-19 depends on the age and health conditions of each patient. Children are not considered in the present study because they rarely develop symptoms upon contracting COVID-19. We assign an age to each individual agent. For this purpose, we use a distribution representing the age structure of a population of sampled agents. The age of the agents determine the risk of COVID-19-related mortality as shown in Table 4 [6]. Each agent can also have one or many of the following risk factors: cardiovascular diseases, blood pressure, diabetes, cancer, and chronic respiratory diseases. The prevalence of each of these risk factors and the corresponding death probability are given in Table 5 [7]. One of the core assumptions of the model is that we consider the maximum among the probabilities of death caused by the age and the risk factors. For example, if a patient is aged 82 and has diabetes, the death probability will be 14.8% which corresponds to the rate of mortality by old age and not because of diabetes.

**Table 4:** The distribution of age groups in the model and the corresponding mortality risk. Mortality risks are taken from [6].

| Age group | Sampling probability in the model | Death probability |
| --- | --- | --- |
| 18-29 | 19.24 % | 0.2 % |
| 30-39 | 19.24 % | 0.2 % |
| 40-49 | 19.24 % | 0.4 % |
| 50-59 | 19.24 % | 1.3 % |
| 60-69 | 14.57 % | 3.6 % |
| 70-79 | 6.08 % | 8 % |
| 80-90 | 2.39 % | 14.8 % |

**Table 5:**
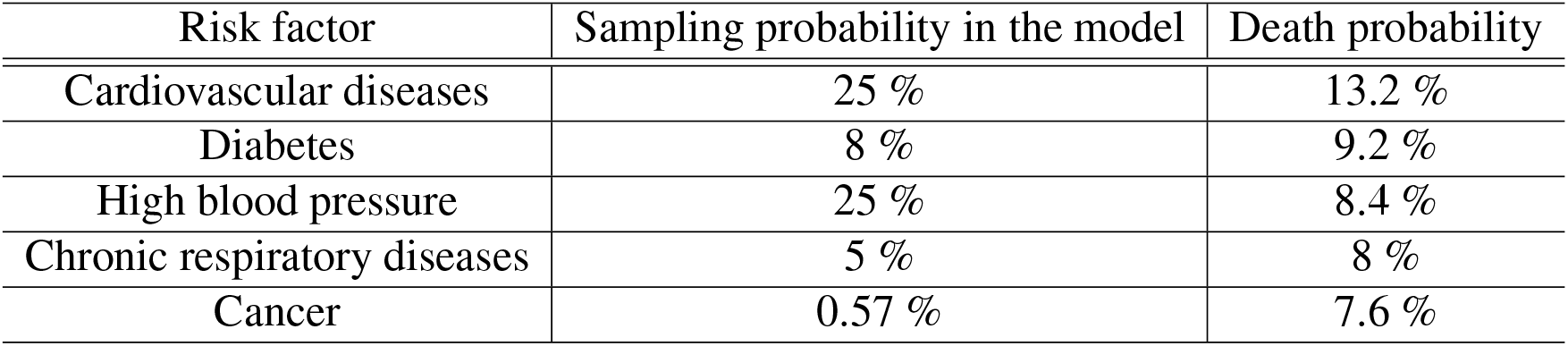
The prevalence of risk factors in the model and the considered mortality rate for each one. Mortality risks are taken from [7].

### 2.5. Computer implementation

The code was written in C++ in the object-oriented programming (OOP) style. The GNU Scientific Library (GSL) was used for the sampling of random variables. The average CPU time for numerical simulations of transmission dynamics in 90 days is around 3.5 hours on a computer with four cores and 6 Gb of RAM. Results were post-processed using the software ParaView and Python scripts. The code is available and can be accessed at: https://github.com/MPS7/SIM-CoV.

## 3. Results

### 3.1. Transmission dynamics in the case of relaxed control measures

We begin by considering the case of low-dense population (*ρ* = 1 *Ped.m^−^*^2^). The socio-psychological force is not taken into account as we do not consider social distancing. We begin by validating the model by considering a case where NPIs are relaxed. To do this, we reproduce data of COVID-19 outbreak in Calabria, Italy [40] by fitting the probabilities of direct and indirect transmission in the model (*p_d_* and *p_in_)*. The values of transmission probabilities which give a good agreement are *p_d_* = 0.04 and *λ* = 0.002. The number of COVID-19 tests in this region is estimated to be around 7500 tests. Figure 4 shows a comparison of the simulated number of cases and the reported ones in Calabria after model calibration. The model displays a similar growth factor as the reported one in data of active infected cases in Calabria, as well as the other regions of Italy.

**Figure 4:**
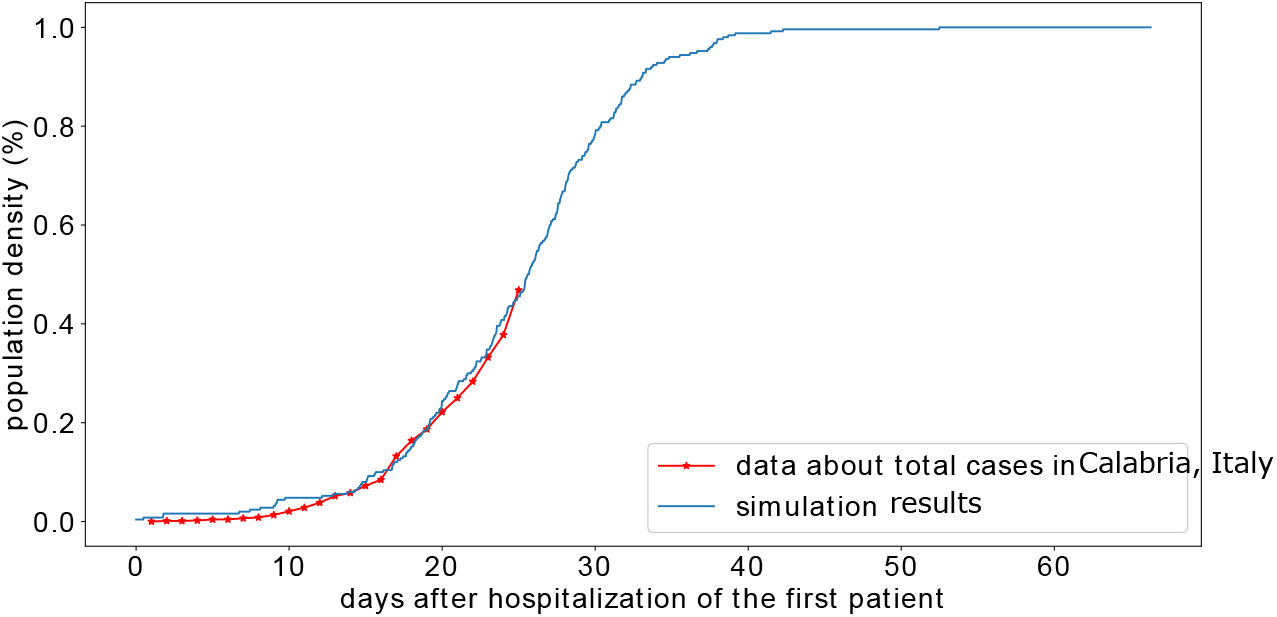
Simulated cumulative proportion of symptomatic cases versus the cumulative fraction of reported cases to the number of tests in the region of Calabria, Italy [40].

After model calibration, we study the dynamics of disease transmission under relaxed epidemic control measures. We run a numerical simulation and we study the evolution of agent populations. The mean age of the sampled population is 45.05 and 53 % of sampled individuals have at least one of the five risk factors considered in the model. The simulation starts by introducing one infected agent to the computational domain. The exponential growth of infected agents begins approximately 14 days after that. In this phase, the cumulative number of infected agents doubles in approximately every four days. We have represented the evolution of the active and cumulative fraction of infected agents in Figure 7, A.

**Figure 5:**
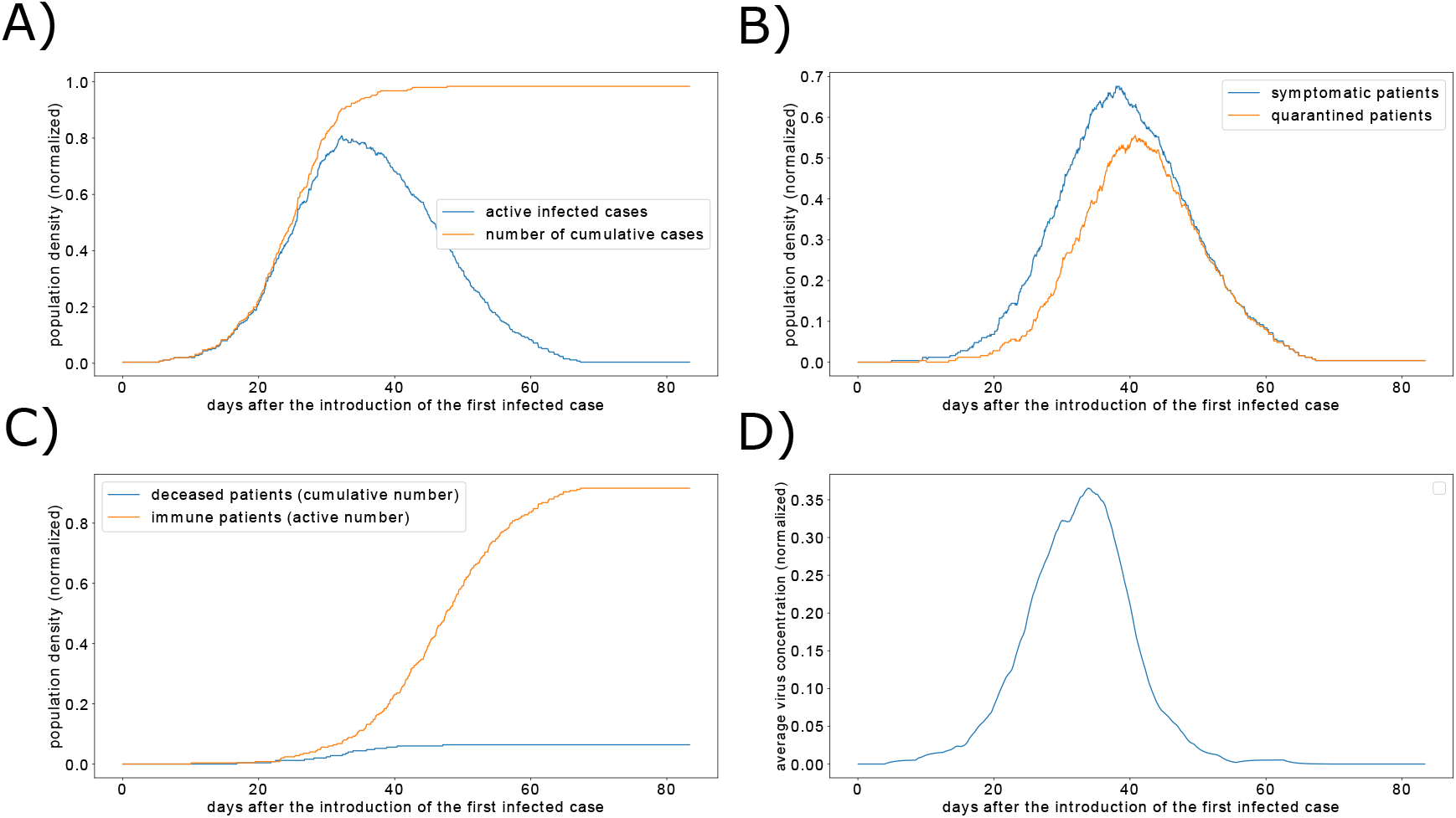
Simulated dynamics of COVID-19 outbreak in the absence of control measures. A) The active and cumulative fraction of infected agents. B) The populations of symptomatic and quarantined patients over time. C) The proportions of immune and deceased agents. D) The average SARS-CoV-2 concentration in the computational domain.

The peak of the outbreak is reached in day 31. At this time, the fraction of simultaneously active infected agents reached 80 %. 64 % of them are symptomatic and 37 % are quarantined (Figure 7, B). The outbreak is resolved by day 67. At this time, only one quarantined patient remains and 91.6 % of agents are immunized as shown in Figure 7, C. The model predicts that, in the absence of any control intervention, the disease will infect 98.4 % of the system population by the of the simulation time. The model predicts that the case fatality rate (CFR) is 6.5 %. Note that these values can slightly change from one simulation to another because of the stochastic noises caused by the relatively low number of agents. Snapshots of numerical simulations showing the different stages of the epidemic are provided in Figure 6. The concentration of stable SARS-CoV-2 on hard surfaces reaches its maximal value during the peak of the outbreak. This suggests that indirect tranmission become more frequent as the diseases propagates.

**Figure 6:**
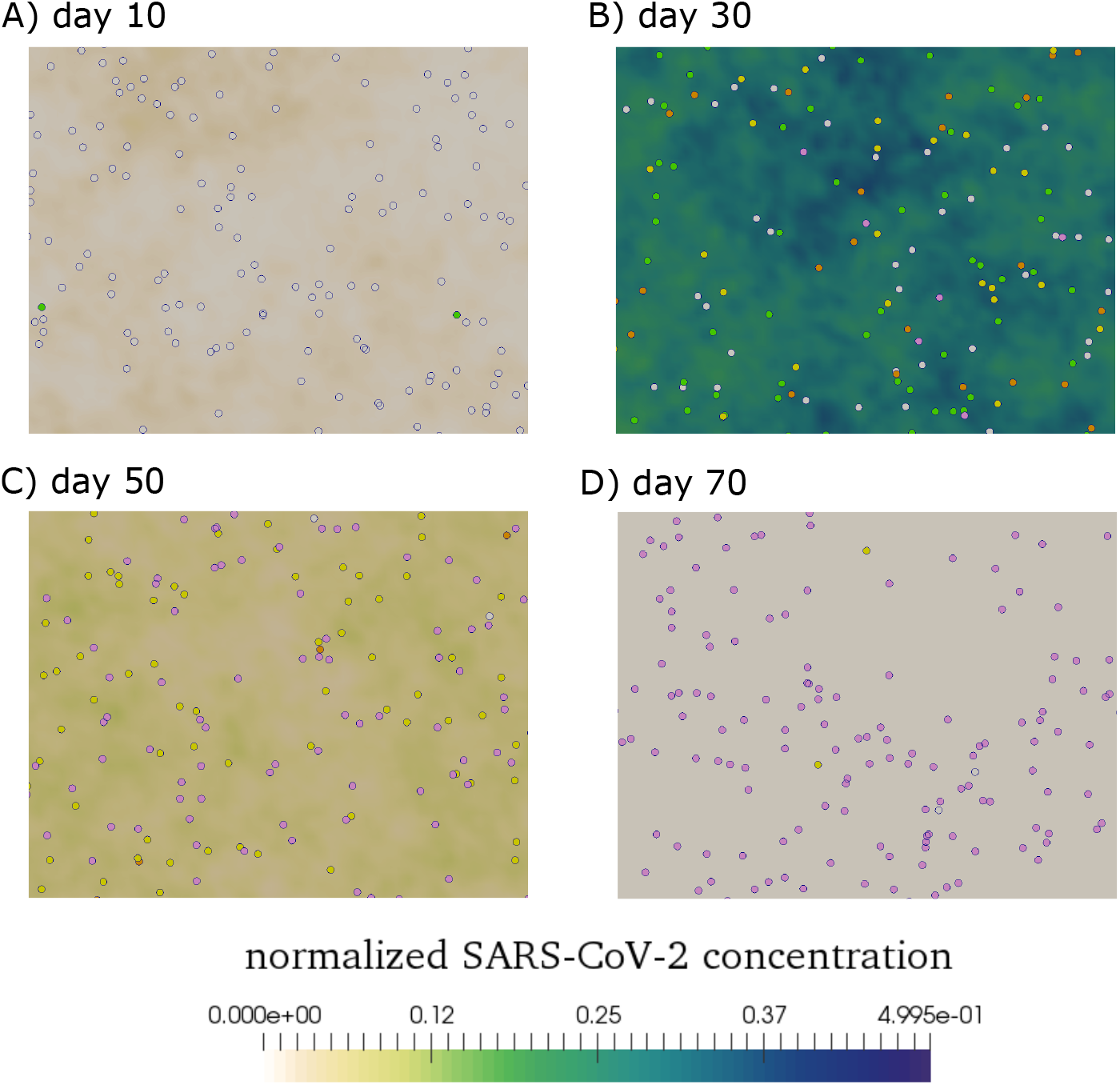
Screenshot of a numerical simulation showing four moments of the transmission dynamics after the introduction of one infected agent to the system. The legend of agent types is given in Figure 1. A) Before the onset of the infection, only a few agents are infective and transmit the disease to susceptible agents. B) The peak of disease propagation, most of the agents are infected. Some are symptomatic while others are quarantined. The virus concentration in the computational domain reaches its maximal value during this time. C) The number of mobile infective individuals starts to decrease, most of the agents are either quarantined or immunized. D) The epidemic is over and most of the agents are now immunized and cannot become infected again.

### 3.2. Panic situations increase disease transmission in crowded areas despite social distancing

Social distancing, or physical distancing, is a set of non-pharmaceutical interventions which aim to increase the minimal distance between people. Social-distancing is effective in slowing down the epidemic progress because it prevents contacts between individuals. However, it is hard to accurately estimate that minimal distance that should be kept between individuals to avoid contacts in crowds and, especially in panic situations. Indeed, keeping a distance from everyone becomes hard when an individual is moving inside a crowd of panicking people. To assess the risk of infection spread in crowds, we run numerical simulations of high-density crowds and we estimate the frequency of person-to-person contact. First, we study the effect of density on crowd dynamics in the absence of the socio-psychological force. We define a contact as the situation where the distance between the two agents is smaller to the sum of their radius. Figure 7, A shows the exponential growth of the contact frequency as the agent density increases. Then, we introduce the socio-pyschological force and we study the impact of its magnitude (*α*) on the contact frequency in the crowd. The magnitude of the socio-psychological force quantifies the tendency of individuals to comply with social distancing measures. Figure 7, B illustrates the evolution of the contact frequency as a function of the magnitude of the socio-psychological force for two density values. As expected, the contact frequency decreases as the magnitude grows. However, this frequency can still be important for average magnitudes of the socio-psychological force, in high-density crowds.

**Figure 7:**
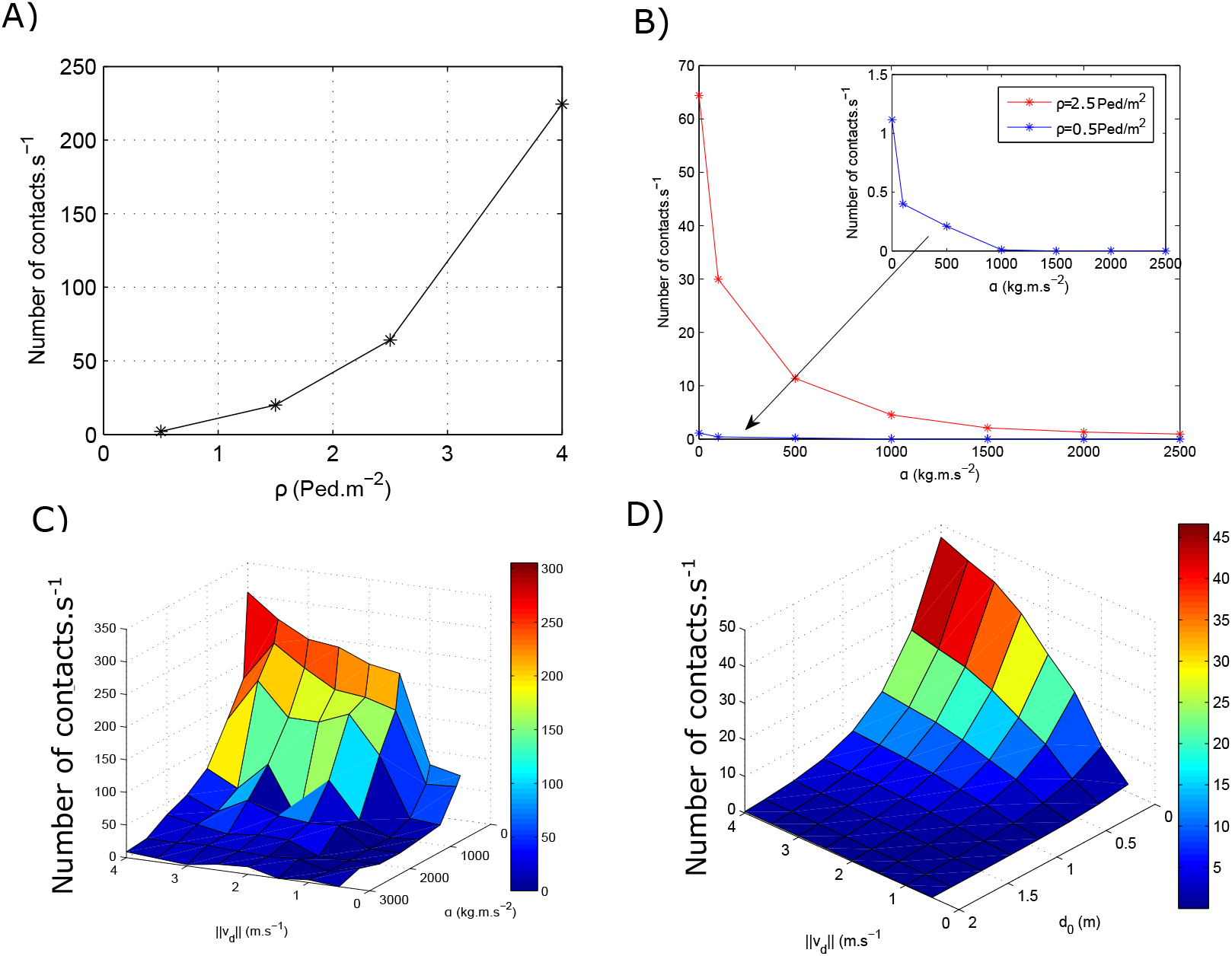
High-density crowds and panic situations increase the risk of person-to-person tranmission. A) Contact frequency as a function of agent density *ρ*. B) Contact frequency versus the amplitude of socio-psychological force *α*. C) Frequency of contacts versus the amplitude of socio-psychological force *α* and the speed of desired velocity ||*v_d_*||. D) Contact frequency as a function of the desired distance *d_0_* and the speed of desired velocity ||*v_d_*|.

In normal situations, the amplitude of the desired velocity ||*v_d_*|| is sampled from a normal distribution with an average of 1.34 *m.s^−^*^1^ and a standard deviation equal to 0.26 *m.s^−^*^1^. Usually, agents keep the desired minimal distance between them if they walk at this speed in uncrowded areas. On the other hand, contact frequency increases significantly when agents are under panic (||*v*_d_|| > 3 *m.s^−^*^1^) or if they move in a dense crowd (*ρ* ≥ 1.5 *Ped.m^−^*^2^). In this case, the number of contacts can be minimized only if the magnitude of the socio-psychological force is very high. Figure 7, C shows the contact frequency as a function of both amplitudes of the desired velocity and the socio-psychological force in a crowd with a density equal to 1.5 *Ped*.*m^−^*^2^. These findings suggest that individuals will no longer keep the minimal distance between them as a safety measure when they are under panic. This scenario was observed in train and metro stations (for example in Italy and in India) when a large number of people attempted to travel a few days before the start of the curfew. Therefore, it is important to think about other control measures such as the use of restricted access and obstacles to reduce the flow of people.

Next, we consider a fixed value for the magnitude of the socio-psychological force, taken equal to *α* = 2000 *kg.m.s^−^*^2^ and we change the desired distance that agents aim to keep between them (do). In Figure 7, D, we represent the contact frequency as a function of the amplitude of the desired speed and the distance that agents wish to keep between them (*d*_0_). As before, we notice the contact frequency decreases when the desired distance is higher than 1.5 *m*. This indicates that the measures which aim to maintain a distance of one metre between individuals may not be sufficient to prevent contacts in dense crowds, under panic.

### 3.3. Pre-symptomatic transmission significantly accelerates the onset of the exponential epidemic growth

In the remainder of this paper, we study the effect of transmission dynamics in uncrowded areas with a density equal to 1 *Ped*.*m*−^2^. We do not consider the socio-psychological force as we no longer consider social distancing. Several studies have suggested the possibility of pre-symptomatic transmission in COVID-19 [5, 41, 42]. However, it is not clear if this pre-symptomatic transmission significantly affects the dynamics of the epidemic. To investigate this hypothesis, We suppose that infected individuals start transmitting the disease before the onset of symptoms. We run three simulations reflecting different periods of pre-symptomatic transmission: *h* (no pre-symptomatic transmission), 24 *h*, and 48*h*. Simulations predict that adding twenty-four hours to the period of pre-symptomatic transmission accelerates the onset of the exponential growth by approximately five days. Decreasing the period of pre-symptomatic period also flattens the epidemic curve of COVID-19 as shown in Figure 8. This suggests that the early diagnostic for COVID-19 will flatten the epidemic curve. The isolation of pre-symptomatic carriers should significantly slow down the spread of the disease.

**Figure 8:**
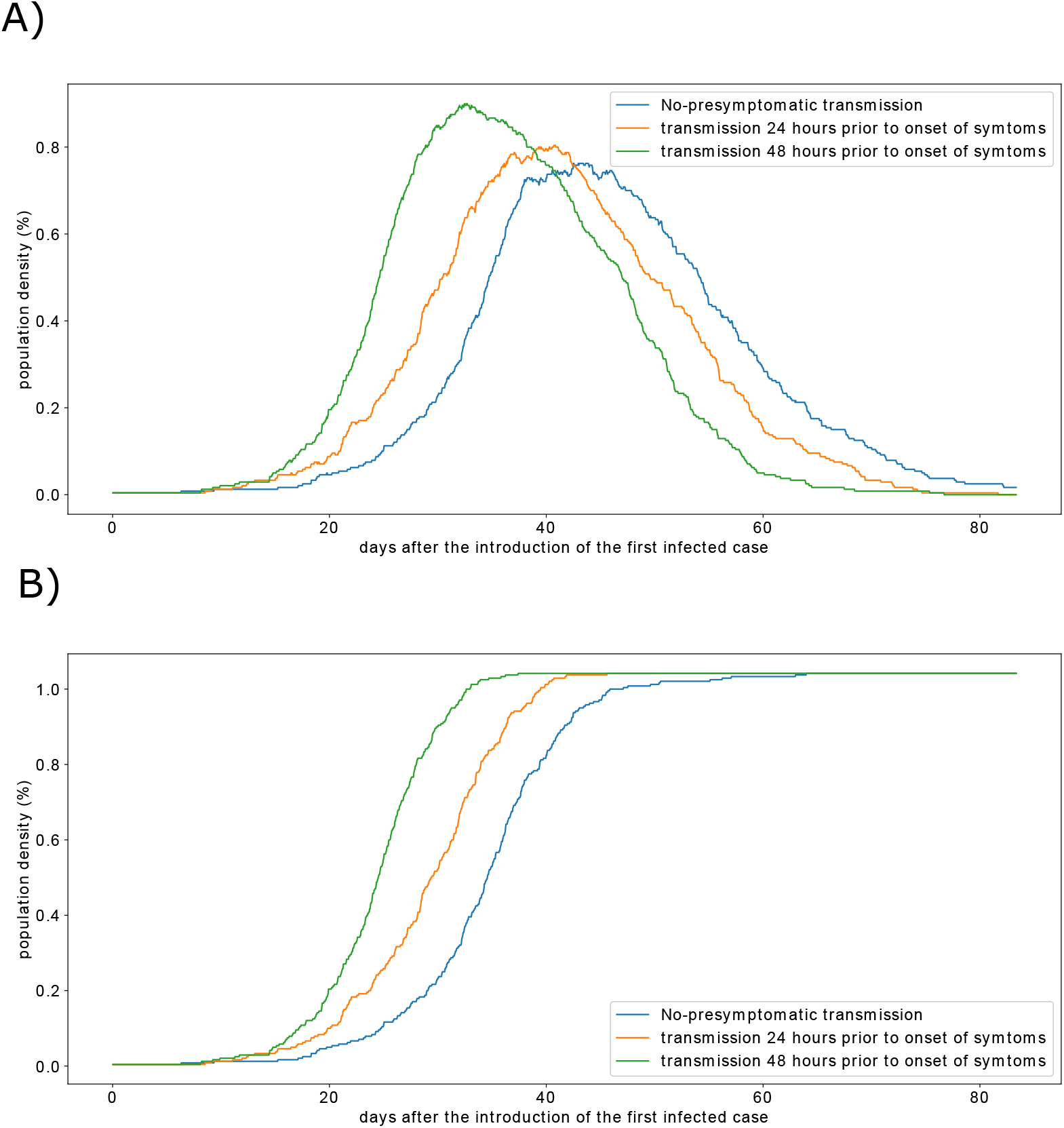
The effect of the duration of pre-symptomatic transmission on active infected cases (A) and the cumulative proportion of infected individuals (B).

### 3.4. SARS-CoV-2 persistence on the hard surfaces increases the number of infected individuals reached at the epidemic peak

An important feature of SARS-CoV-2 is its ability to persist for a few hours to a few days on hard surfaces. In fact, SARS-CoV-2 is more stable on plastic and stainless steel than on copper and cardboard [3]. The half-life time of the virus varies from 2 hours to 10 hours depending on the surface type. To evaluate the effect of SARS-COV-2 persistence on its transmission dynamics, we compare the output of three numerical simulations. We run each simulation with the same parameter set except for the SARS-CoV-2 decay rate (σ) which corresponds to three values of SARS-CoV-2 half-life times on hard surfaces: 2 *h*, 4 *h*, and 8 *h*. The results show that the epidemic dynamics remain the same except for the value of infected individuals reached during the peak of the outbreak (Figure 9).

**Figure 9:**
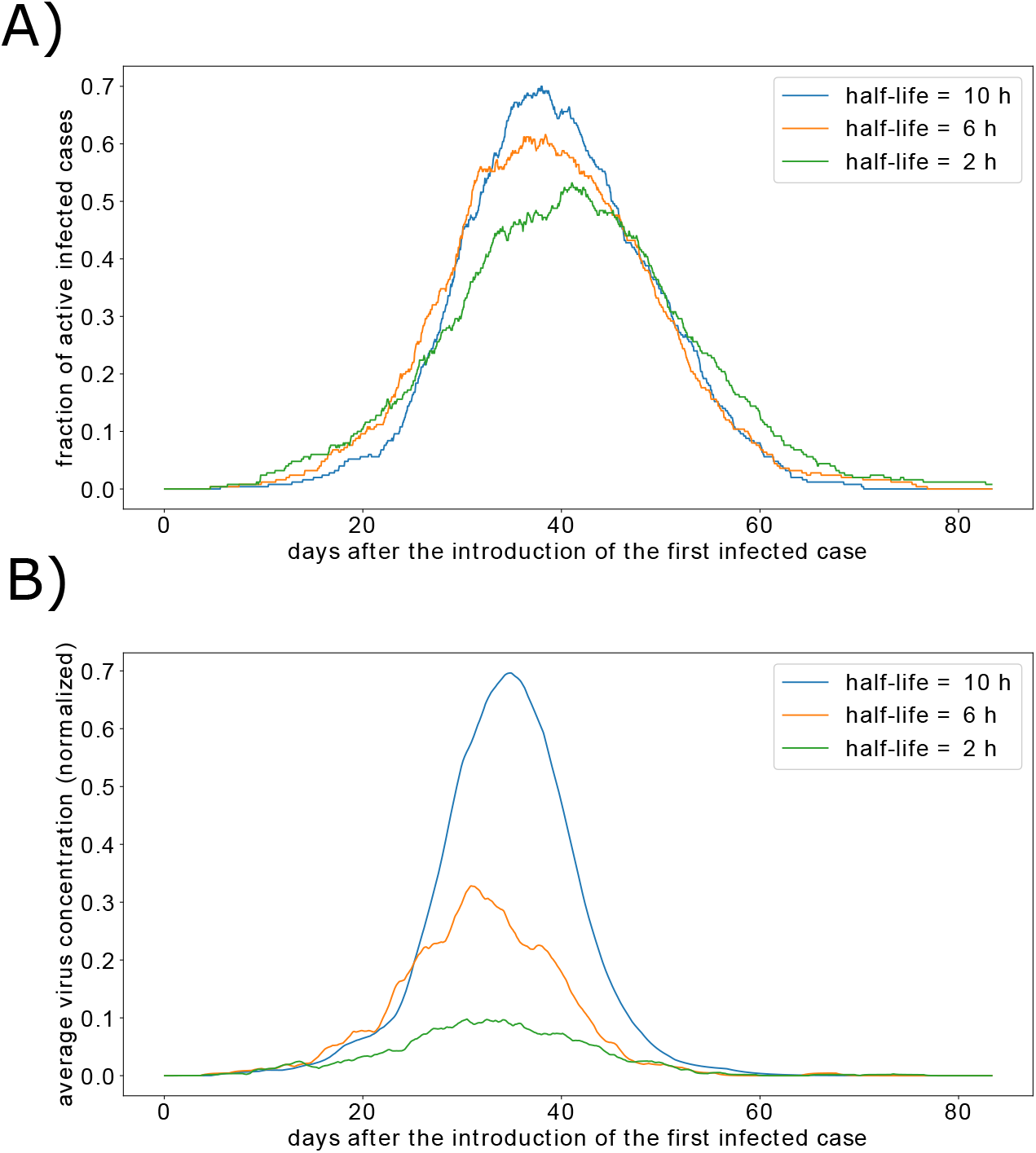
Transmission dynamics for three different constants of SARS-CoV-2 decay. A) The fraction of active infected agents over time. B) The average concentration of the virus in the computational domain.

To further investigate this effect, we count the number of direct and indirect transmissions in each simulation. In the normal case, where the half-life time is 4 h, the percent of indirect transmissions was 23 %. Most of these transmissions took place during the peak of the outbreak because the virus concentration is high at this moment. Increasing the persistence of the virus resulted in a higher proportion of indirect transmission to 28.1 *%*. While reducing the half-life time of the stable virus decreases the rate of indirect transmission to 19.8 %. Disinfection of public areas reduces the concentration of stable SARS-CoV-2 on hard surfaces. Therefore, we expect it to decrease the number of indirect transmissions and to reduce the reached number of infected individuals during the outbreak peak.

### 3.5. Restricting the movement of the population flattens the epidemic curve of COVID-19

Lockdown is an emergency control measure which aims to reduce the movement of the total or a part of the population. It can be used to mitigate the spread of highly infections diseases such as COVID-19. However, a full lockdown is a costly solution as it causes the arrest of all economic activities. We investigate the effect of partial lockdown on disease spread by considering that only a proportion of the population can move. The model predicts that higher the number of motionless agents is, the flatter the epidemic curve will be (Figure 10, A). In other words, the slope of the exponential growth decreases as the number of immobile agents grows. The cumulative fraction of infected patients is also reduced as shown in Figure 10, B. Restricted population movement also decreases the proportion of infected agents (Figure 10, C) and the number of infected individuals reached during the peak of the outbreak (Figure 10, D). As a result, partial lockdown measures reduce the number of deceased patients as shown in Figure 10, E.

**Figure 10:**
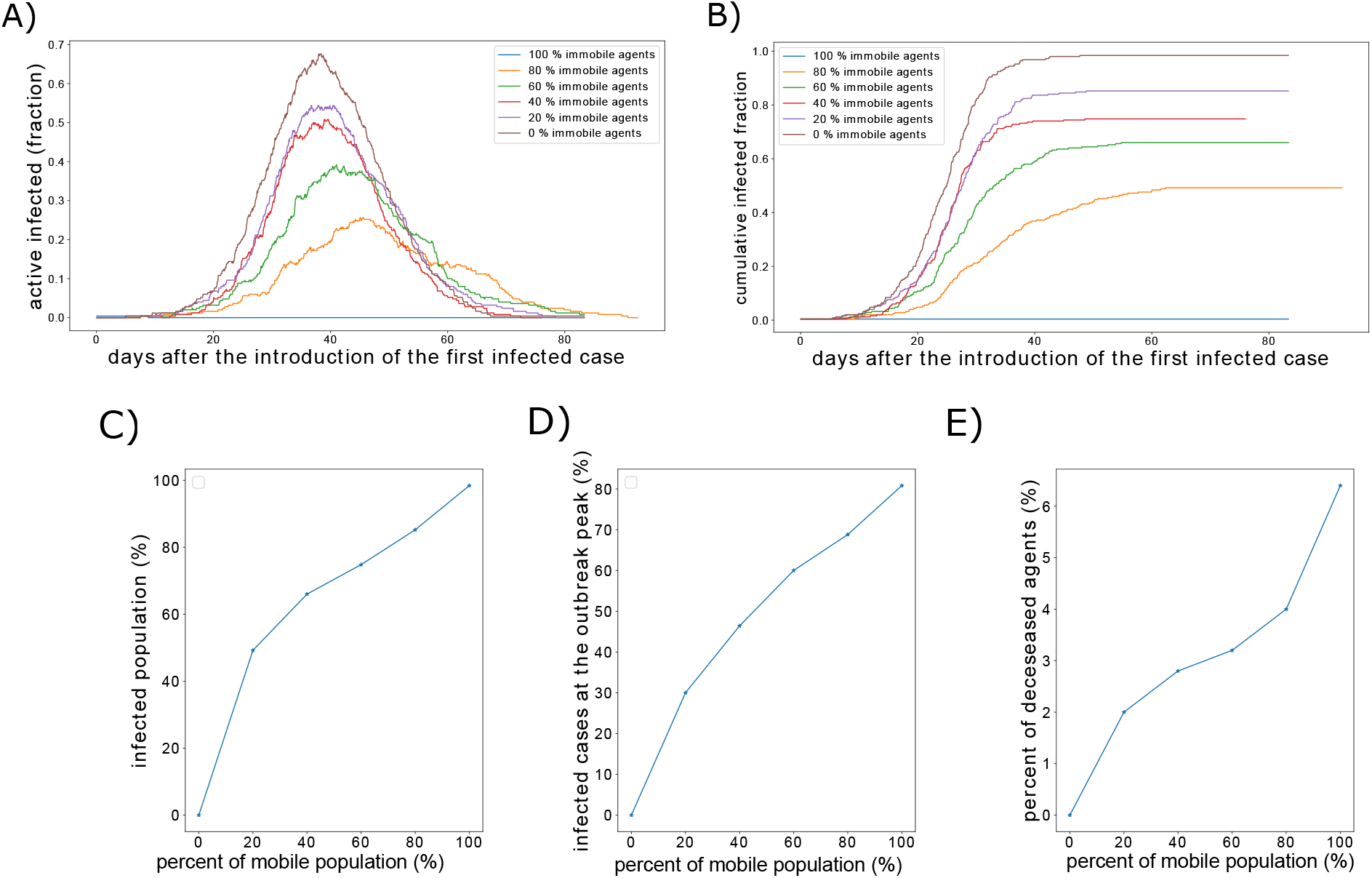
The effect of restricted population movement on the epidemic dynamics of COVID-19 outbreak. A) Populations of infected individuals. B) Cumulative fraction of infected individuals. C) The total infected proportion of the population as a function of movement restriction level. D) The effect of restricted movement on the percent of infected individuals at the peak of the outbreak. E) The proportion of deceased individuals versus the percent of immobile agents.

We compare the model predictions with the reported data of the cumulative number of hospitalized patients in Wuhan, China [43]. To achieve this, we fit both the percent of immobile agents and other model parameters. We notice that reducing the percent of mobile agents flattens the epidemic curve. However, it does not delay the onset of the exponential growth in a similar way to the reported epidemic curve for the case of Wuhan, China. Therefore, we have reduced the probabilities of transmission by approximately four times to observe roughly the same epidemic curve (Figure 11).

**Figure 11:**
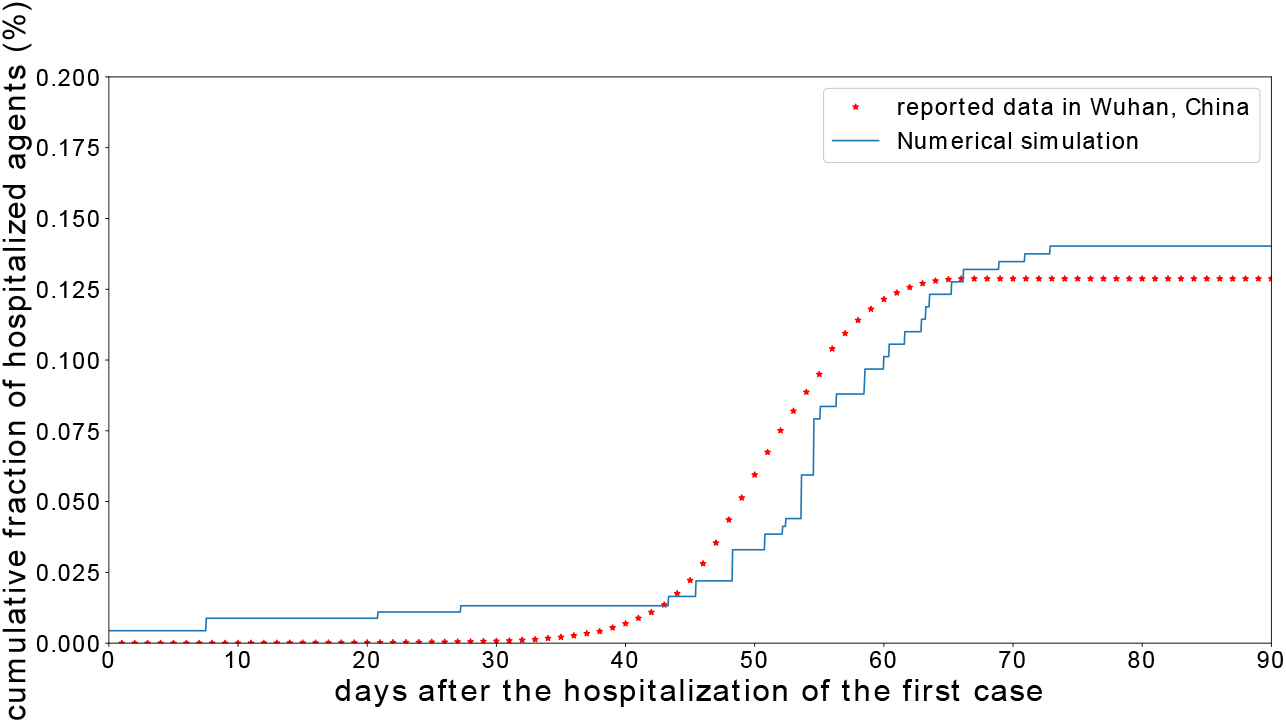
Comparison of the simulation result for restricted population movement and low transmission rate, with data reported in Wuhan, China. The cumulative percent of reported cases in Wuhan, China, was computed by dividing the number of cases by 230000, which corresponds to the number of COVID-19 tests.

## 4. Discussion

This paper is devoted to the study of COVID-19 transmission dynamics using a specific multi-scale model. We calibrated the model using available data on the cumulative number of reported cases in two situations, one for relaxed control measures (Calabria, Italy) and the other with strict non-pharmaceutical interventions (Wuhan, China). The proposed model has several features which makes it suitable for the quantification of the effects of control measures on both the individual and population levels. First, we have used a microscopic social force model to describe the motion of agents. As a result, we were able to properly study the effect of social distancing in crowded areas. Numerical simulations with this model revealed that these measures do not prevent the transmission of the disease in high-density areas, especially when agents move under panic. Next, we considered the mode of indirect disease transmission which depends on the virus concentration on contaminated surfaces. This transmission mode is important in COVID-19 as SARS-CoV-2 can remain viable on various surfaces for a few days [3].

Another important feature of the model concerns the integration of measured characteristics of the clinical course of COVID-19. The incubation period was sampled using a log-normal distribution, which was fitted using clinical data [4]. Mean values for the periods from symptom onset to hospitalization, discharge, and death were taken the literature [39]. Finally, The realistic distributions of demographic and clinical factors allows the accurate prediction of the population-specific CFR. We can tune the model to predict the response of different countries with different age structures and risk factors prevalence to the spread of COVID-19.

Several conclusions can be drawn from the present study concerning the effect of epidemic control interventions. First, we have shown that panic situations could result in the spread of the disease, despite the adherence of individuals to social distancing measures. Numerical simulations with the social force model suggests that social distancing is not fully respected in crowded areas, especially when individuals move under panic. Situations such as panic-buying could result in the rapid spread of the virus. Therefore, control strategies for COVID-19 should aim to avoid the formation of dense crowds and should prevent the spread of panic. Furthermore, we have demonstrated that keeping a minimal distance of one metre between individuals is not sufficient to prevent person-to-person contacts in crowded areas. Next, the model have shown that pre-symptomatic transmission significantly shapes the outbreak dynamics. Therefore, modelling works devoted to the study of COVID-19 spread should take into consideration the effect of pre-symptomatic carriers. Besides, increasing the number of tests for COVID-19 is expected to significantly slow down the propagation of the epidemic as many pre-symptomatic carriers will get quarantined before they transmit the disease. After that, we speculate that the disinfection of public areas would result in a decrease in the number of patients reached during the peak of the outbreak. This is because we observe the same thing when we increase the decay rate of SARS-CoV-2 on hard surfaces. Finally, fitting the model to reproduce epidemiological data on the case of Wuhan, China, suggests that measures that are stricter than social distancing and lockdown were adopted to control the epidemic spread. We can speculate that the early wearing of masks, the washing of hand, and the following of cough etiquette, not only flatten the epidemic curve but also delay the onset of the exponential growth.

Note that the model relies on a few key assumptions. First, the effect of unreported cases and asymptomatic transmission were not considered in the present work. The role of these ‘silent transmitters’ of infectious diseases was investigated in a previous work [44]. We did not introduce them in the present work, as there are no sufficient data on the infection dynamics of COVID-19 in asymptomatic individuals. We will study the effect of unreported cases using our model in a forthcoming work. Second, the model considers a relatively low number of agents. This is because of the high computational cost of the model. We have used periodic boundary conditions to approximate systems with a higher number of individuals. We expect the results to be similar for larger systems but with less stochastic noises. Finally, the study is devoted to the transmission dynamics in closed systems. The transport of people between the studied area and other locations could impact the epidemic dynamics. In the future, we will apply the model to investigate the transmission dynamics of COVID-19 in crowds, in urban areas with realistic configurations.

## Data Availability

The entire set of equations and parameter values used is presented in the article. The used code to generate the results can be obtained at: https://github.com/MPS7/SIM-CoV

